# Emergence and widespread circulation of a recombinant SARS-CoV-2 lineage in North America

**DOI:** 10.1101/2021.11.19.21266601

**Authors:** Bernardo Gutierrez, Hugo G. Castelán Sánchez, Darlan da Silva Candido, Ben Jackson, Shay Fleishon, Christopher Ruis, Luis Delaye, Andrew Rambaut, Oliver G. Pybus, Marina Escalera-Zamudio

## Abstract

Genetic recombination is an important driving force of coronavirus evolution. While some degree of virus recombination has been reported during the COVID-19 pandemic, previously detected recombinant lineages of SARS-CoV-2 have shown limited circulation and been observed only in restricted areas. Prompted by reports of unusual genetic similarities among several Pango lineages detected mainly in North and Central America, we present a detailed phylogenetic analysis of four SARS-CoV-2 lineages (B.1.627, B.1.628, B.1.631 and B.1.634) in order to investigate the possibility of virus recombination among them. Two of these lineages, B.1.628 and B.1.631, are split into two distinct clusters (here named *major* and *minor*). Our phylogenetic and recombination analyses of these lineages find well-supported phylogenetic differences between the Orf1ab region and the rest of the genome (S protein and remaining reading frames). The lineages also contain several deletions in the NSP6, Orf3a and S proteins that can augment reconstruction of reliable evolutionary histories. By reconciling the deletions and phylogenetic data, we conclude that the B.1.628 major cluster originated from a recombination event between a B.1.631 major virus and a lineage B.1.634 virus. This scenario inferred from genetic data is supported by the spatial and temporal distribution of the three lineages, which all co-circulated in the USA and Mexico during 2021, suggesting this region is where the recombination event took place. We therefore support the designation of the B.1.628 major cluster as recombinant lineage XB in the Pango nomenclature. The widespread circulation of lineage XB across multiple countries over a longer timespan than the previously designated recombinant XA lineage raises important questions regarding the role and potential effects of recombination on the evolution of SARS-CoV-2 during the ongoing COVID-19 pandemic.

## Introduction

Virus recombination, the process by which genetic material from two genetically distinct parental lineages is combined into a viable descendant virus genome, is a common feature of sarbecovirus evolution (Boni et al, 2020). Genomic analyses suggest that recombination events among coronaviruses circulating in non-human species occurred during the evolutionary history of SARS-CoV-2 (Zhu et al, 2020; Li et al, 2020; Lytras et al, 2021). Signals of ongoing recombination among SARS-CoV-2 genomes assessed under a statistical framework during the COVID-19 pandemic have been reported (VanInsberghe et al., 2021). Notably, SARS-CoV-2 genomes that are clearly recombinant have been observed at low frequencies in the UK, some of which showed evidence of forward transmission (Jackson et al, 2021). One of these UK recombinants was designated as lineage XA, the first recombinant lineage in the Pango nomenclature system (O’Toole et al., 2021; Rambaut et al., 2020).

Our understanding of the effects of genomic recombination on SARS-CoV-2 fitness and transmission dynamics are still limited, but genetic exchange has been previously associated with evolutionary adaptation in viruses under experimental conditions (e.g. Poliovirus; Xiao et al., 2016), in individual hosts (e.g. HIV; Song et al., 2018) and in nature (e.g. human influenza viruses; Petrova & Russell, 2018). Interestingly, a recombination event is associated with the emergence of a MERS-CoV lineage that became dominant in camels in the Middle East between 2014 and 2015 (Sabir et al., 2016). Although few clearly recombinant SARS-CoV-2 lineages have been reported so far, our ability to detect them is likely to increase as time progresses, due to the continued genetic divergence of SARS-CoV-2 and increased co-circulation of divergent lineages. Thus, the question regarding the potential for recombination to contribute to SARS-CoV-2 evolution and adaptation remains.

For virus recombination to occur, the parental lineages need to co-circulate in the same location in order to allow specific individuals to become co-infected. This scenario provides the circumstances during which chimeric genotypes can emerge, usually through molecular processes such as template switching, homologous recombination or reassortment (the latter occurring in viruses with segmented genomes; Simon-Loriere & Holmes, 2011). Coronaviruses naturally produce a variety of recombination products during natural infection, including recombinant genomes, a process mediated by the proofreading exoribonuclease (Gribble et al., 2021).

Mosaic genomes resulting from recombination can be detected through changes in sequence similarities among different regions of the virus genome relative to parental lineages. Identifying recombination between recently diverged lineages is difficult, because sequence similarities are high, and it is hard to distinguish homoplasic changes from those that are identical by descent due to inheritance from a recent shared ancestor (synapomorphic changes). In such instances, other mutations such as insertions and deletions can prove informative; specifically, deletions are highly unlikely to revert during the evolution of a single lineage. Phylogenetic methods can also provide a tractable approach to test hypotheses regarding virus recombination, as they can be used to reconstruct the separate evolutionary histories of subgenomic regions (Simon-Loriere & Holmes, 2011). While genome regions that share the same ancestry can be easily established for segmented viruses (e.g., Orthomyxoviruses, such as influenza viruses [Holmes et al, 2005]), the exact start and endpoints of recombinant genome regions must be inferred statistically for non-segmented viruses (Pérez-Losada et al, 2015). Furthermore, estimating the timing and location of recombination events can be limited by uncertainty in estimates of phylogenetic node ages, although such uncertainty can be reduced by using methods that combine evolutionary information across different genome regions (e.g., Raghwani et al, 2012).

As SARS-CoV-2 circulates around the world, new lineages emerge and are tracked using the Pango dynamic hierarchical nomenclature system (Rambaut et al, 2020). During late 2020 and early 2021, a series of lineages descending from B.1 were first detected in North and Central America. Specifically, lineages B.1.627, B.1.628, B.1.631 and B.1.634 were detected by the national genomic surveillance programs in the United States of America, Mexico and other countries in the Americas and their genomes shared publicly on the GISAID database (Shu & McCauley, 2017). Genomic similarities among these and other lineages circulating in the region and elsewhere prompted the suggestion that recombination had occurred during their emergence and spread (this hypothesis was initially proposed and discussed on the Pango GitHub website: https://github.com/cov-lineages/pango-designation/issues/189).

Here, we present an analysis of the spread and evolution of these four lineages and investigate the possibility that one or more recombination events contributed to their evolution. Our results provide evidence supporting a recombinant origin for lineage B.1.628 and its designation as a distinct recombinant lineage that presented forward transmission, circulating in multiple countries.

## Methods

### Genomic data, metadata and sequence alignment

We retrieved all complete SARS-CoV-2 genome sequences assigned to Pango lineages B.1.627, B.1.628, B.1.631 and B.1.634 as of August 30 2021 from GISAID (Shu & McCauley, 2017). Accompanying sequence metadata, including sampling locations (at different geographic resolutions) and dates of sample collection and submission were also retrieved. This complete data set included 1950 sequences that were subsequently filtered to exclude all sequences for which >10% of sites were ambiguous (i.e., had nucleotide states N or X). The final data set, comprising 1055 genome sequences, was used for all phylogenetic analyses. The original complete data set (n=1950) was used in part to explore the spatio-temporal distribution of the four Pango lineages under investigation.

After adding the reference SARS-CoV-2 genome Hu-1 (GenBank accession MN908947.3; Wu et al 2020) to the filtered data sets, the sequences were aligned using MAFFT v7.487 (Katoh & Standley, 2013). The resulting alignments were inspected visually to identify deletions >1nt in length and which were shared by all or most of the sequences assigned to one or more of the lineages under investigation; these deletions were removed from the alignment and encoded as discrete sequence traits, which were later mapped onto estimated phylogenetic trees (see *Results*).

### Confirmation of Pango lineage assignment and whole genome phylogenetic analysis

The Pango lineage assignment of the sequences in our final data set was determined using Pangolin v.3.1.11 (Rambaut et al., 2020); all lineages originally assigned on GISAID were confirmed. To further explore the phylogenetic structure of our data, we constructed a maximum likelihood (ML) phylogenetic tree using IQtree (Minh et al., 2020) under a GTR+Γ substitution model. We estimated node support values using the Shimodaira-Hasegawa (SH) approximate Likelihood Ratio test (SH-aLRT; Guindon et al., 2010), with 1000 replicates, and 1000 bootstrap replicates.

### Genome-wide divergence of Pango lineages and recombination breakpoint inference

Pairwise genetic distances across the length of the genome of basal sequences for each of the lineages under investigation and the reference Wuhan-Hu-1 genome were estimated using custom scripts (available at https://github.com/BernardoGG/XB_lineage_investigation), based on the ape package in R (Paradis & Schliep, 2019). One sequence was selected per lineage, and in cases where multiple important clades were identified within single Pango lineage one basal sequence from each of these clades was included. Raw genome-wide distances were estimated for 500-nucleotide segments, and a sliding window approach was used to estimate these distances across segments that overlapped every 20 nucleotides.

Recombination tests are computationally demanding for large data sets. To address this, we further subsampled the alignment to include one sequence per country per Pango lineage per day (script available at https://github.com/BernardoGG/XB_lineage_investigation), resulting in a downsized set of 716 whole genome sequences. To evaluate recombination patterns in our data, we further reduced the downsized data set by randomly sampling 200 sequences from the four lineages under investigation. This reduced data set was then analysed using GARD (Kosakovsky Pond et al, 2006a), a tool that uses a genetic algorithm to search for one or more putative breakpoints in a multiple sequence alignment; the best supported number of non-recombinant fragments in the alignment is then evaluated by comparing the Akaike Information Criterion (AIC_c_) of the proposed models versus a null model (i.e., no recombination points, such that a single tree topology best explains the sequence alignment). The phylogenetic topological incongruencies of potential non-recombinant fragments can be evaluated to identify lineages involved in the recombination event (Kosakovsky Pond et al, 2006b).

### Phylogenetic analyses of inferred non-recombinant genome segments

Given the possibility that a single phylogeny of the complete genome alignment does not explain the evolutionary history of our sequences, we partitioned the alignment using the inferred breakpoints from GARD (derived from the analysis of the 200-sequence data set). ML trees for each genome partition were then estimated with IQtree as described previously.

We identified four key deletions amongst our sequences under investigation: two deletions in the NSP6 gene (Orf1ab) and two deletions in Orf3a (see *Results*). NSP6 deletions occur at two adjacent locations (here called ΔR1 and ΔR2) and don’t result in changes in the reading frame. Orf3a deletions occur on a single locus and take the form of either a single-nucleotide frameshift deletion (ΔFS2) or a 4-nucleotide frameshift deletion (ΔFS4). These deletions were coded as discrete characters, assigned to individual tree tips and reconstructed at internal nodes using a parsimony criterion, thereby visualising the history of their occurrence across the phylogenies; these patterns have to reconcile with the proposed recombination events. Our rationale is that the individual evolutionary histories of each of the putative non-recombinant fragments should also parsimoniously explain the occurrence of deletions observed in NSP6, S and ORF3a, under the assumption that deletions do not revert once they occur in a lineage. Our rationale also draws from the premise that these deletions are more likely to descend from single occurrences within the evolution of each lineage but are not restricted to have occurred just once across the whole phylogeny. Specifically, some of these deletions have been observed previously in other lineages and variants of concern, including the NSP6 deletions observed in B.1.1.7, P.1 and B.1.351 (Meng et al., 2021). The genomic position where a given deletion occurs (relative to the inferred recombination breakpoints) was used to determine which genome partition most likely represents the true evolutionary history of that deletion. Loci where the deletion was flanked by ambiguities were differentially labelled with an asterisk (*).

### Exploring the phylogenetic discrepancies of the lineages under investigation relative to other B.1 lineages

Our phylogenetic analyses showed that lineages B.1.628 and B.1.631 are split into two groups each. B.1.628 contains a cluster of sequences that fall near the root of the phylogenies and are henceforth identified as *B*.*1*.*628 minor*, and a large more derived monophyletic clade henceforth identified as *B*.*1*.*628 major*. B.1.631 is split into a small cluster of sequences that consistently cluster near the tree backbone in both genome segments and is henceforth identified as *B*.*1*.*631 minor*, and a larger monophyletic clade here called *B*.*1*.*631 major*. To explore the topological discrepancies between the lineages under investigation in the absence of B.1.631 minor (see *Results* for an explanation of why this cluster was excluded), we randomly sampled five sequences from each of the lineages under investigation (B.1.627, B.1.628 major, B.1.628 minor, B.1.631 major and B.1.634), with five random sequences from the B.1.1.7 lineage (i.e. Alpha variant of concern, VOC) and five random sequences from the B.1.351 lineage (i.e. Beta VOC); this was performed in order to explore the congruency of the diversification patterns of the lineages under investigation in context of the B.1 lineage. Five additional random sequences from lineage A.2.5 were also included to represent the Pango A lineage and to provide an outgroup for tree rooting. All sequences from the B.1.1.7, B.1.351 and A.2.5 lineages were obtained from GISAID; they had sampling dates that spanned the time when these lineages were observed to circulate and were predominantly from locations in North and Central America. We performed GARD analyses and constructed ML phylogenetic trees from this data set as previously described. We estimated node support for the phylogenetic analyses using 1000 bootstrap replicates, and nodes with >50% node support were collapsed into polytomies.

Finally, we used the snipit software (https://github.com/aineniamh/snipit) to explore the distribution of single nucleotide polymorphisms (SNPs) across the genome of potential recombinant and parental lineages. SNPs were identified and visualised in reference to the Wuhan Hu-1 genome sequence.

## Results

### Distribution of lineages B.1.627, B.1.628, B.1.631 and B.1.634

We noted the spatiotemporal distribution of a total of 1950 sequences for lineages B.1.627 (n = 252), B.1.628 (n = 1391), B.1.631 (n = 181) and B.1.634 (n = 126). Sequences from the four lineages were collected between July 8 2020 and August 18 2021 (Table 1), however the majority were sampled in 2021 (Fig 1A). All four lineages were predominantly sampled in North America (89.5% of sequences), either in the United States of America (USA) or Mexico. B.1.627 and B.1.631 were mostly sampled in the USA whereas B.1.634 was commonly found in Mexico (Fig 1B). Lineage B.1.628 is the most geographically widespread lineage in our data set: it has been identified in 41 different USA states and in 31 Mexico states (all other lineages were identified in 10-21 USA states and in 15-17 Mexico states; Fig 1C). B.1.628 is also the most widely sampled through time, with 406 days between the earliest and most recent sample collection date (compared to B.1.627 = 212 days, B.1.631 = 232 days, B.1.634 = 160 days). B.1.628 was sampled in the USA during the entirety of this sampling period, while only sampled for a period of 185 days in Mexico.

**Table 1.**
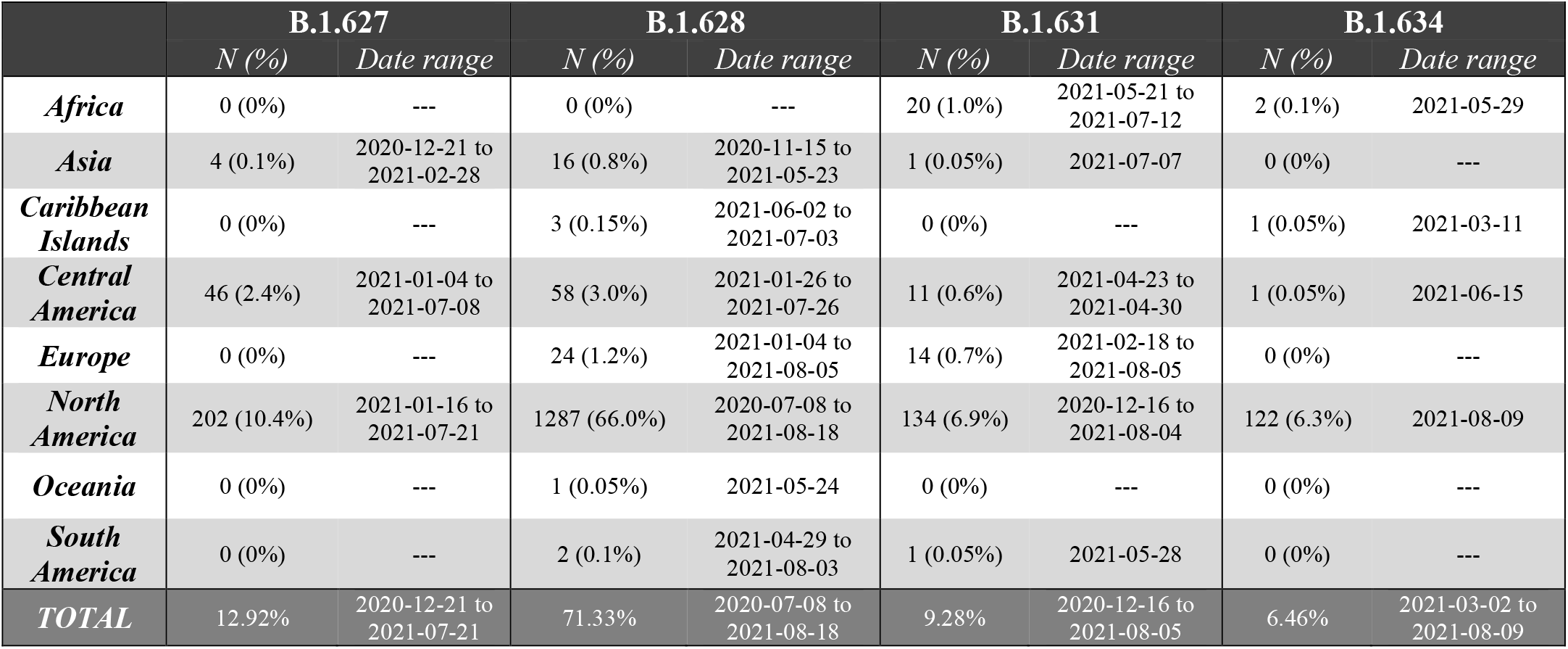
Summary of sequences from four SARS-CoV-2 lineages associated with potential recombination (percentages shown in reference to the complete number of sequences in our data set).

|  | B.1.627 |  | B.1.628 |  | B.1.631 |  | B.1.634 |  |
| --- | --- | --- | --- | --- | --- | --- | --- | --- |
|  | *N (%)* | *Date range* | *N (%)* | *Date range* | *N (%)* | *Date range* | *N (%)* | *Date range* |
| *Africa* | 0 (0%) | --- | 0 (0%) | --- | 20 (1.0%) | 2021-05-21 to 2021-07-12 | 2 (0.1%) | 2021-05-29 |
| *Asia* | 4 (0.1%) | 2020-12-21 to 2021-02-28 | 16 (0.8%) | 2020-11-15 to 2021-05-23 | 1 (0.05%) | 2021-07-07 | 0 (0%) | --- |
| *Caribbean Islands* | 0 (0%) | --- | 3 (0.15%) | 2021-06-02 to 2021-07-03 | 0 (0%) | --- | 1 (0.05%) | 2021-03-11 |
| *Central America* | 46 (2.4%) | 2021-01-04 to 2021-07-08 | 58 (3.0%) | 2021-01-26 to 2021-07-26 | 11 (0.6%) | 2021-04-23 to 2021-04-30 | 1 (0.05%) | 2021-06-15 |
| *Europe* | 0 (0%) | --- | 24 (1.2%) | 2021-01-04 to 2021-08-05 | 14 (0.7%) | 2021-02-18 to 2021-08-05 | 0 (0%) | --- |
| *North America* | 202 (10.4%) | 2021-01-16 to 2021-07-21 | 1287 (66.0%) | 2020-07-08 to 2021-08-18 | 134 (6.9%) | 2020-12-16 to 2021-08-04 | 122 (6.3%) | 2021-08-09 |
| *Oceania* | 0 (0%) | --- | 1 (0.05%) | 2021-05-24 | 0 (0%) | --- | 0 (0%) | --- |
| *South America* | 0 (0%) | --- | 2 (0.1%) | 2021-04-29 to 2021-08-03 | 1 (0.05%) | 2021-05-28 | 0 (0%) | --- |
| *TOTAL* | 12.92% | 2020-12-21 to 2021-07-21 | 71.33% | 2020-07-08 to 2021-08-18 | 9.28% | 2020-12-16 to 2021-08-05 | 6.46% | 2021-03-02 to 2021-08-09 |

**Figure 1.**
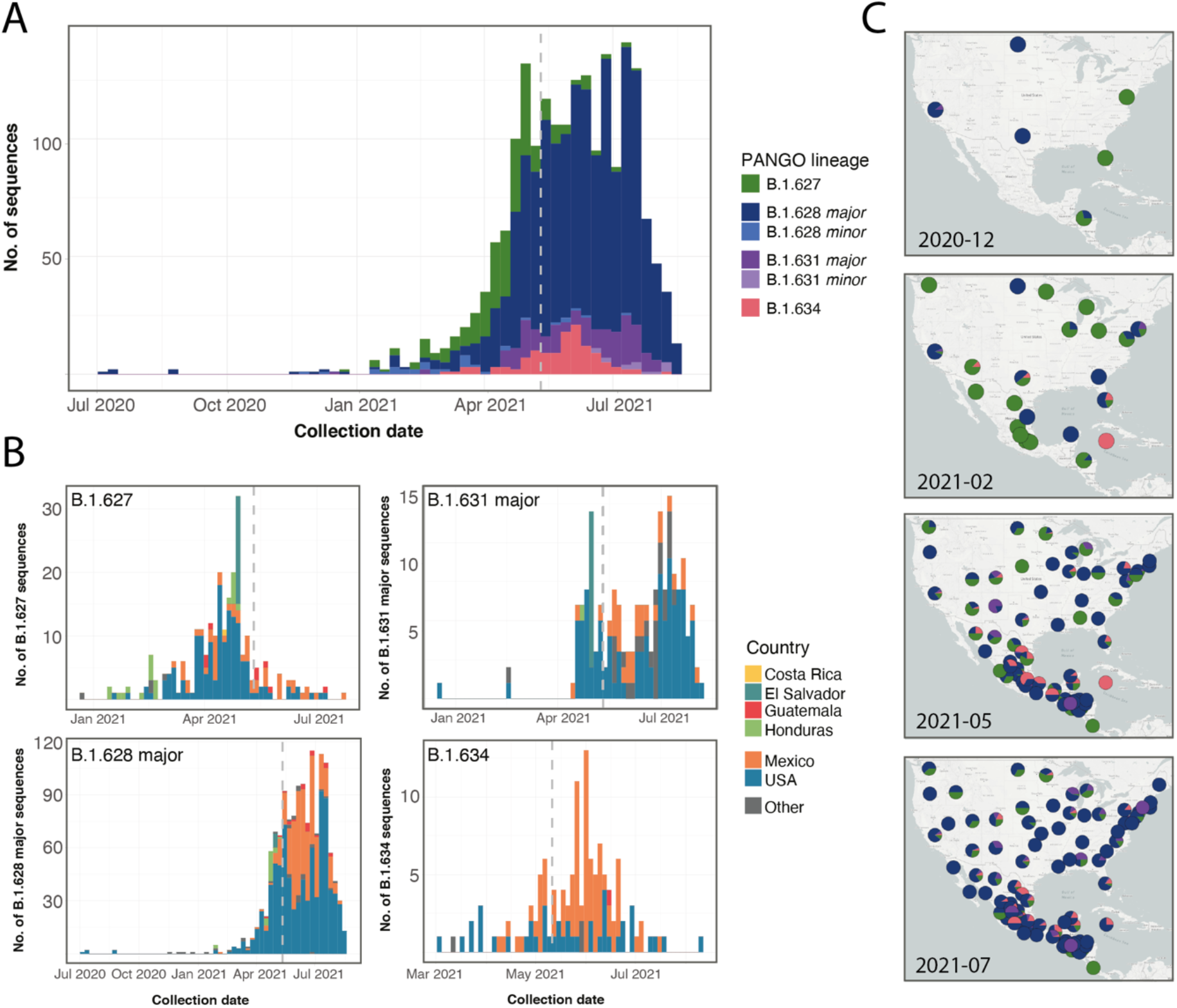
Spatiotemporal distribution of lineages B.1.627, B.1.628 (major and minor), B.1.631 (major and minor) and B.1.634. **(A)** Number of sequences on GISAID (per two-week period, as of 2021-08-30) for each lineage publicly available in GISAID, plotted using the associated sample collection date for each sequence. **(B)** Weekly number of sequences for each lineage coloured by country from where samples were collected. Countries outside of North and Central America make up <5% of sequences and are therefore grouped in the “Other” category. The dotted vertical lines show the starting of the systematic genome sampling and sequencing programme according to the national SARS-CoV-2 genome surveillance program for Mexico (May 11, 2021). B.1.628 minor and B.1.631 minor lineages are not shown. **(C)** Mapping of the geographic spread of the four lineages in North and Central America (a region where >95% of the sequences were identified) at four representative dates over their complete sampling date range.

A maximum likelihood (ML) tree inferred from these genomes and rooted on the reference genome Wuhan Hu-1 shows that all four lineages form monophyletic clusters as expected (Fig. S1). Two exceptions are noted for lineages B.1.628 and B.1.631. For the former, a group of sequences close to the root of the tree are designated as lineage B.1.628 and this group is distinct from the main B.1.628 clade. In the latter case, sequences from lineage B.1.631 are split into two paraphyletic clusters by B.1.627 (Fig S1 inset). For reference purposes within this work, we henceforth refer to the larger monophyletic B.1.628 and B.1.631 clades as *B*.*1*.*628 major* and *B*.*1*.*631 major*, and identify the sequences clustering at the base of the phylogeny that were assigned to these lineages as *B*.*1*.*628 minor* and *B*.*1*.*631 minor*. Some of the nodes that define important splits in the tree show moderate support; for example, the node that groups most B.1.631 genomes with other B.1.627 genomes (to the exclusion of the outlying B.1.631 genomes) is well supported (SH-aLRT = 98.6; Fig S1). The presence of these phylogenetic clusters that don’t match the Pango lineage definitions (Rambaut et al., 2020) warrants further investigation, with recombination as a possible explanatory factor.

### Recombination analyses and breakpoint inference

Our results indicate that recombination is likely to have occurred in our data set. The GARD analysis suggests that a single breakpoint in the alignment can explain the data (Fig 2A), with high support for a model incorporating this recombination event (Δc-AIC_*null model*_ = 202.176). The breakpoint is inferred by GARD to have occurred around position 21308 (a TTT codon), corresponding to the signal peptide region at the N-terminus of the spike protein (18 nucleotides downstream of the canonical sarbecovirus transcription regulatory sequence AACGAAC; Yang et al., 2021). However, some variation in the results is observed when using different subsampling approaches; an analysis that excludes the B.1.634 lineage indicates a recombination breakpoint is inferred position 22775-22778 (at a GAT codon) in the Spike protein reading frame (Δc-AIC_*null model*_ = 562.098), corresponding to amino acid residue 390D located in the core region of the receptor binding domain (RBD) adjacent to beta sheet 3 (β3; Lan et al., 2020). For either scenario, the receptor binding motif (RBM) which includes the main ACE2 receptor contact points would have been inherited from the same parental lineage (B.1.628).

**Figure 2.**
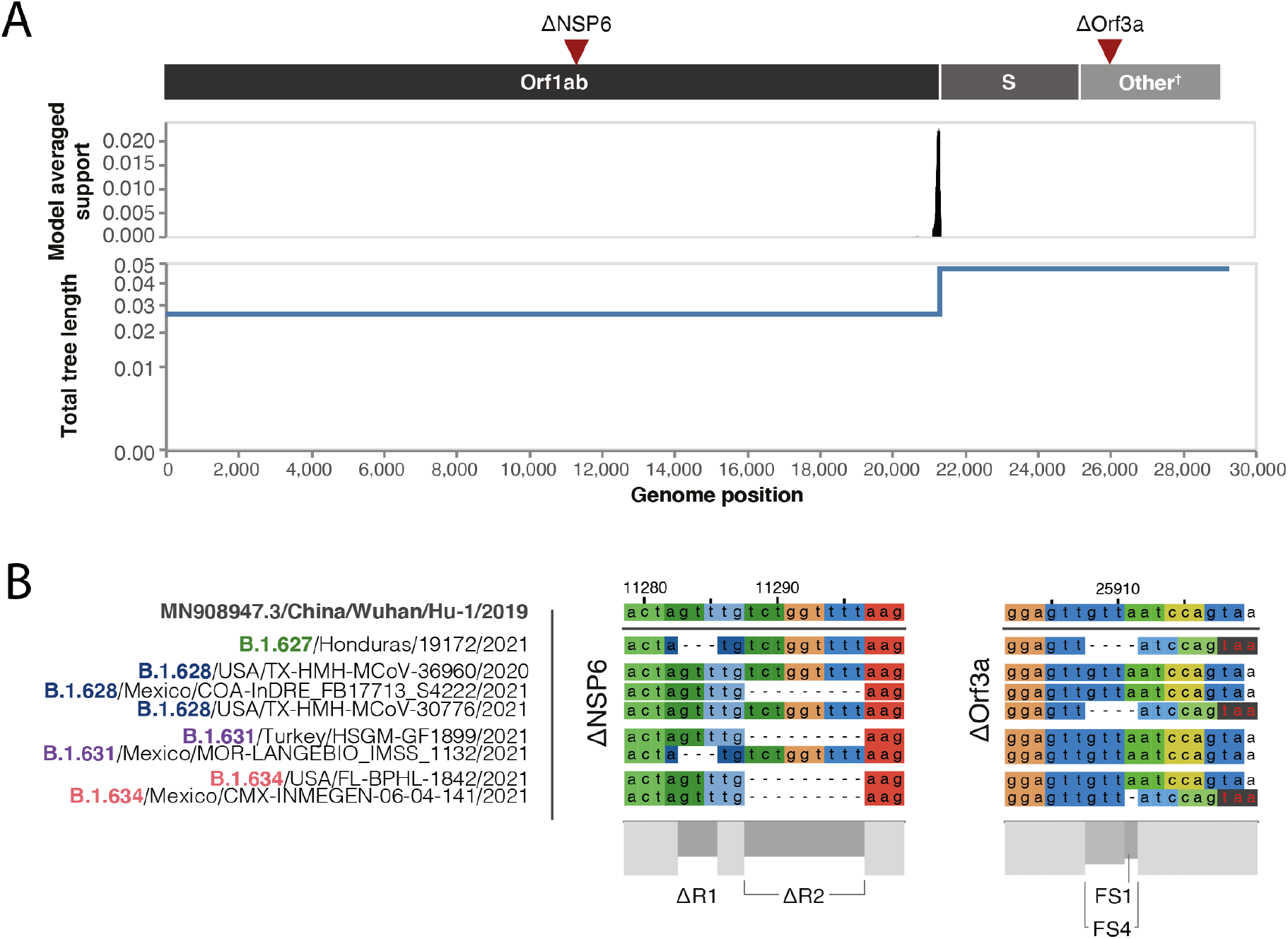
Recombination breakpoint analysis and deletions occurring in the four lineages. **(A)** Recombination breakpoint analysis results performed on GARD shows a statistically supported change in total tree length that stems from an inferred recombination breakpoint around the beginning of the S gene reading frame. The genomic location of deletions under investigated (ΔNSP6 and ΔOrf3a) are shown for reference. **(B)** Deletions in the NSP6 gene (Orf1ab) and Orf3a, illustrated on a representative selection of sequences that includes the B.1.627, B.1.628, B.1.631 and B.1.634 lineages. NSP6 deletions (ΔR1 and ΔR2) and Orf3a deletions (a single-nucleotide frameshift deletion ΔFS2 or a 4-nucleotide frameshift deletion (ΔFS4) are shown, with the early TAA stop codon produced by the Orf3a deletions shown in red letters on a black background.

The recombination analysis outcome also results in the placement of the NSP6 deletions on one side of the breakpoint, and the Orf3a deletions on the other side of the breakpoint (Fig 2A, 2B). The NSP6 region contains two non-overlapping deletions: a 3-nucleotide deletion (ΔR1) and a 6-nucleotide deletion (ΔR2) which are 2 nucleotides apart, but do not derivate in frame shifts downstream (Fig 2B). The Orf3a displays two overlapping frameshift deletions: a single-nucleotide deletion (FS1) and a 4-nucleotide deletion (FS4) which overlap on the fourth position of FS4. Both deletions lead to the same early TAA/UAA termination codon, six nucleotides downstream of the FS1/FS4 locus (Fig 2B).

### Phylogenetic discrepancies between non-recombinant genome segments

Given the inferred recombination breakpoint close to the start of the S reading frame (Fig 2A), we estimated separate phylogenetic trees for Orf1ab and for the remainder of the genome (including S and the remaining structural and non-structural genes, henceforth referred to as the S-3’ region). The phylogenies show topological discrepancies which coincide mostly with individual Pango lineages (Fig 3). Both phylogenies (rooted on the Hu-1 reference genome) show a poorly resolved early split, with Orf1ab showing a bifurcation into two monophyletic groups containingB.1.628 minor and B.1.631 minor basal sequences. On the other hand, the phylogeny for the S-3’ region places the B.1.628 minor sequences as a paraphyletic group from which B.1.634 descends (SH-aLRT = 91.9), whilst B.1.631 minor is a predecessor of the B.1.627, the B.1.628 major and the B.1.631 major lineages (SH-aLRT = 79.7). On the Orf1ab phylogeny, lineages B.1.627 and B.1.631 major share a common ancestor (SH-aLRT = 86.5), while the S-3’ tree shows them as being paraphyletic. Lineage B.1.634 is consistently inferred as monophyletic in both trees; in the Orf1ab tree it descends from B.1.631 minor, and in the S-3’ tree from B.1.628 minor. The nodes defining lineages are generally well supported (SH-aLRT > 70.0), except for the basal nodes for the early bifurcation in both trees; statistical support within each lineage showed a combination of unsupported short branches (SH-aLRT = 0.0) and nodes with high support values (SH-aLRT > 75.0).

**Figure 3.**
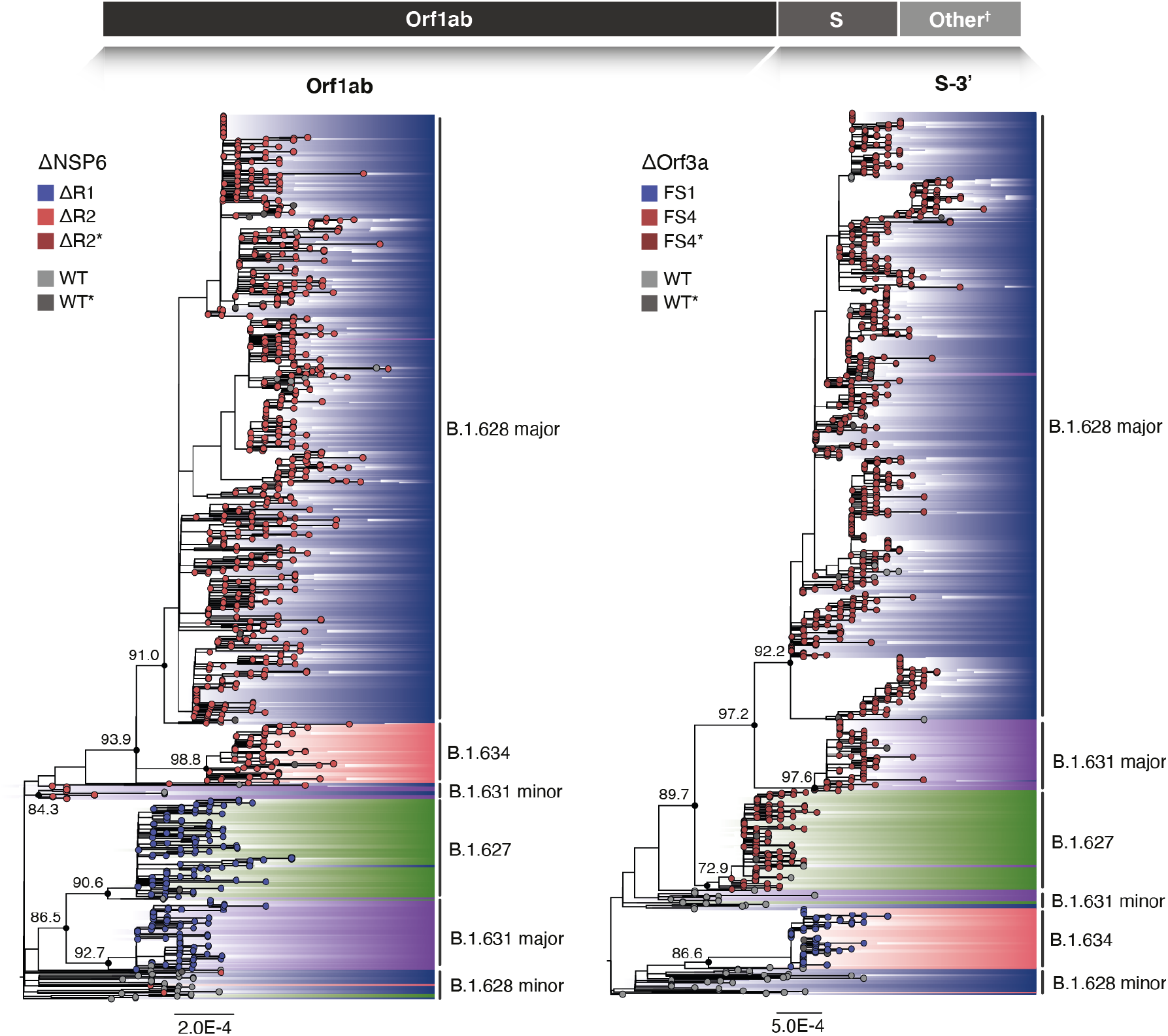
Maximum likelihood phylogenies of two segments of the SARS-CoV-2 genome for the four lineages. Individual phylogenies were constructed for the 5’ and 3’ end of the genome, split at the inferred breakpoint, resulting in trees that represent the evolutionary histories of Orf1ab (left panel) and the S gene plus the remaining structural and non-structural genes† (here referred to as S-3’, right panel). The individually designated Pango lineage for each sequence is highlighted (the consensus identifier for each clade is also shown), and SH-aLRT node support is shown for key lineage defining nodes on both phylogenies. Deletions occurring on each genome segment (ΔNSP6 on Orf1ab and ΔOrf3a on S-3’) are mapped onto the tips of the trees. †Other genes: S, E, M, N, Orf3a, Orf3b, Orf6, Orf7a, Orf7b, Orf8, Orf9b, Orf9c and Orf10.

### Genome-wide divergence across genomes

To further explore the genetic divergence of these lineages and the sequences placed near the root of the trees (specifically, B.1.631 minor and B.1.628 minor), we estimated the pairwise genetic distances across the genomes of representative sequences (basal to the main clades) in reference to the Hu-1 reference genome (Fig S2). While mutations have accumulated in all lineages, the Orf1ab region of B.1.631 minor shows the lowest divergence from Hu-1; all clades display peak genetic divergence between positions ∼21000 and ∼23000, with the exception of B.1.628 major which diverges from Hu-1 homogeneously across its genome.

### Emergence of lineages from a B.1 background and recombination history

Given the genome-wide divergence observed for B.1.631 minor (particularly the unusually high similarities to Hu-1 in the Orf1ab region) and its limited spatiotemporal distribution (i.e. all sequences are from Turkey as opposed to the majority of the sequences that were observed in North and Central America), we excluded this group from further analyses. In particular, the fact that it was not observed anywhere in the Americas suggests that it would not have circulated in the same geographical region, a necessary condition for recombination to occur. In the absence of this cluster, we explored the evolutionary history of the remaining lineages in relation to other lineages that descend from B.1. A phylogenetic analysis including a sample from each lineage under investigation, B.1.1.7 (VOC Alpha) and B.1.351 (VOC Beta) consistently shows that B.1.627, B.1.628 major, B.1.631 major, and B.1.634 group into well-supported monophyletic groups (bootstrap support >93%) for both the Orf1ab (Fig S3) and the S-3’ (Fig S4) genome segments, similar to the well-established Alpha and Beta VOCs. The B.1.628 minor sequences emerge from the polytomy that makes up the B.1 backbone and do not group with the B.1.628 major cluster, suggesting the former do not belong to the B.1.628 Pango lineage. The Orf1ab phylogeny shows that B.1.627 and B.1.631 major share a common ancestor (bootstrap support = 62%), similar to B.1.628 major and B.1.634 (bootstrap support = 100%; Fig S3); this pattern is also observed in the full phylogeny (Fig 3). The S-3’ tree shows that B.1.628 major and B.1.631 major share a common ancestor (bootstrap support = 95%) which in turn descend from a common ancestor with B.1.627 (bootstrap support = 99%), while B.1.634 emerges independently from the B.1 background (Fig S4); once again, the pattern is mirrored by the full S-3’ phylogeny (Fig 3).

Reconciling the occurrence of the NSP6 and Orf3a deletions with the reconstruction of the evolutionary histories of these lineages for both genome segments is possible, as none of the NSP6 deletions occurred simultaneously on the same sequence, and the overlapping nature of the Orf3a deletions (i.e., FS1 is contained within FS4) make it possible to encode ΔR1, ΔR2, FS1 and FS4 as unique traits and to map them to the phylogenetic trees (Fig 3). A third deletion, Δ69-70 on the S protein (also observed in previous VOCs and VOIs; REF**), was exclusive to B.1.634 and therefore not used as an informative marker. From the Orf1ab tree, ΔR1 is shared between B.1.627 and B.1.631 major, while ΔR2 is shared by B.1.631 minor, B.1.628 major and B.1.634. The most parsimonious explanation for this deletion pattern would suggest that ΔR1 occurred once (and predates the ancestral form of the Orf1ab of the B.1.627 and the B.1.631 major lineages) and that ΔR2 occurred once (predating the ancestral form of the Orf1ab for the B.1.628 major and B.1.634 lineages). Similarly, the two distinct frameshift deletions in Orf3a appear to have occurred independently: FS1 occurred once in the ancestral form of lineage B.1.634, and FS4 occurred at least once in the ancestral form of B.1.631 major, B.1.628 major and B.1.627 (Fig 3; Fig 4, upper panel). A considerable number of B.1.628 minor sequences in both trees share the “wild-type” trait (i.e., no deletion) with the Hu-1 reference genome, further suggesting that this group of sequences belongs to either B.1 or a different B.1.X lineage.

**Figure 4.**
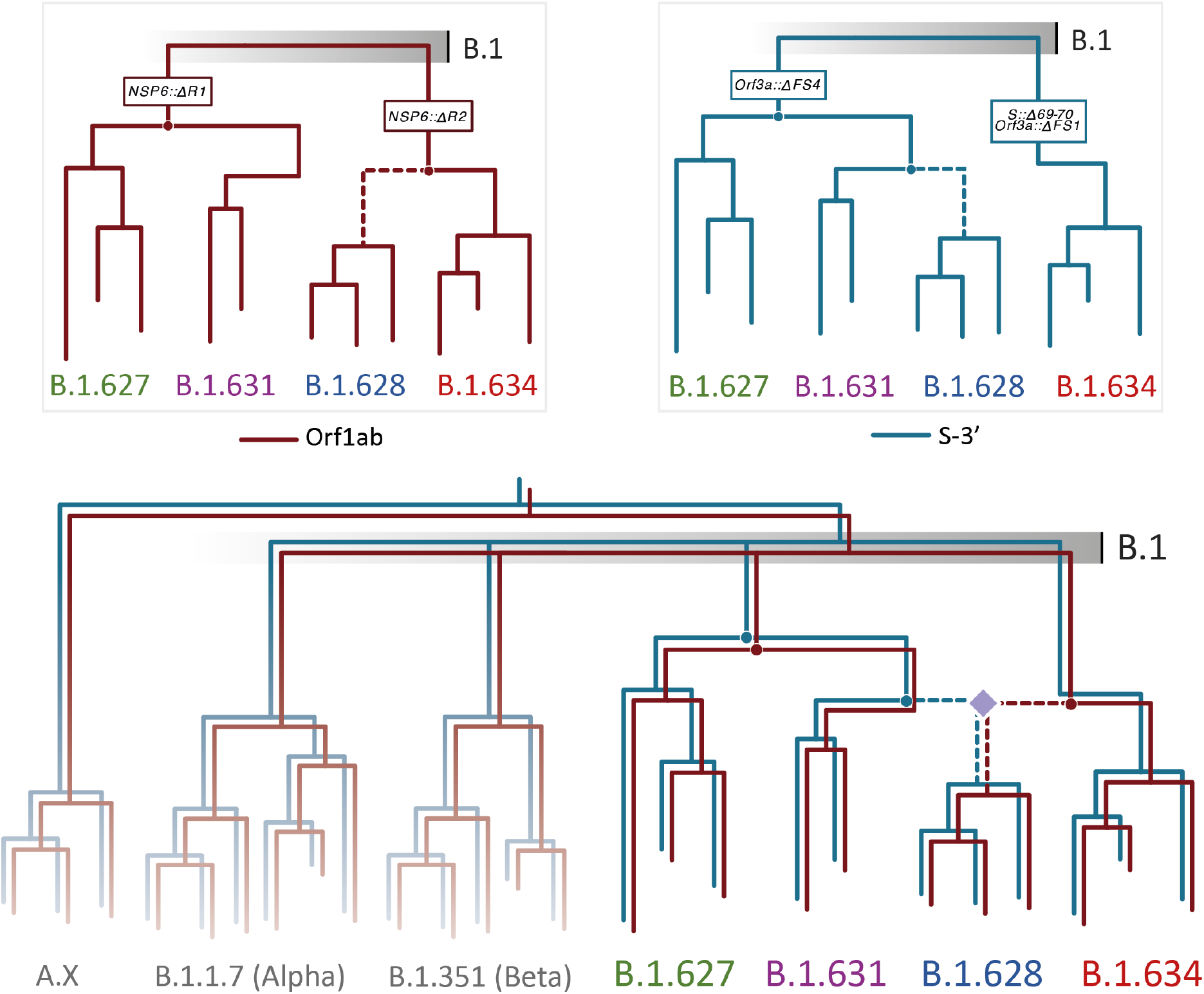
Schematic of the emergence of lineages B.1.627, B.1.628 major, B.1.631 major and B.1.634 from a B.1 background and their recombination history. The phylogenetic reconstruction of the evolutionary trajectories of both genome segments for the four lineages require a single occurrence of each of the deletions in ΔNSP6 and ΔOrf3a to explain most parsimoniously the observed deletion patterns (upper panel). A recombination history that reconciles these deletions while maintaining the inferred ancestors through phylogenetic analyses requires the occurrence of one recombination event, where lineages B.1.631 major and B.1.634 result in the emergence of a recombinant B.1.628 major lineage (lower panel). B.1.627 is evolutionarily related to B.1.631 major but not involved in the recombination event.

Mapping the deletions to a phylogenetic tree inferred for the whole genome and for the complete data set results in these appearing as homoplasic events that require repeated occurrence; specifically, FS4 would have had to occur twice (once in the branch leading to B.1.627 and B.1.631 major and once in B.1.628 major; Fig S5). Therefore, the most parsimonious model that reconciles the minimum required number of deletions and the phylogenetic incongruencies between the evolutionary histories of both non-recombinant genome segments results in B.1.628 major having descended from a recombinant event. It inherited the Orf1ab segment (carrying the ΔR1 deletion on NSP6) from the lineage leading to B.1.634, and the S-3’ segment (carrying the FS4 deletion on Orf3a) from the lineage leading to B.1.631 major (Fig 4). Visualising the SNPs of these lineages shows that B.1.628 major shares at least 6 polymorphisms with B.1.631 major in the first ∼17000 nucleotides of the genome and at least 9 polymorphisms with B.1.634 in the final ∼8000 nucleotides of the genome; no polymorphisms are shared between B.1.628 major and B.1.631 major along the 3’ end of the genome, while no polymorphisms are shared between B.1.628 major and B.1.634 along the 5’ end (Fig 5).

**Figure 5.**
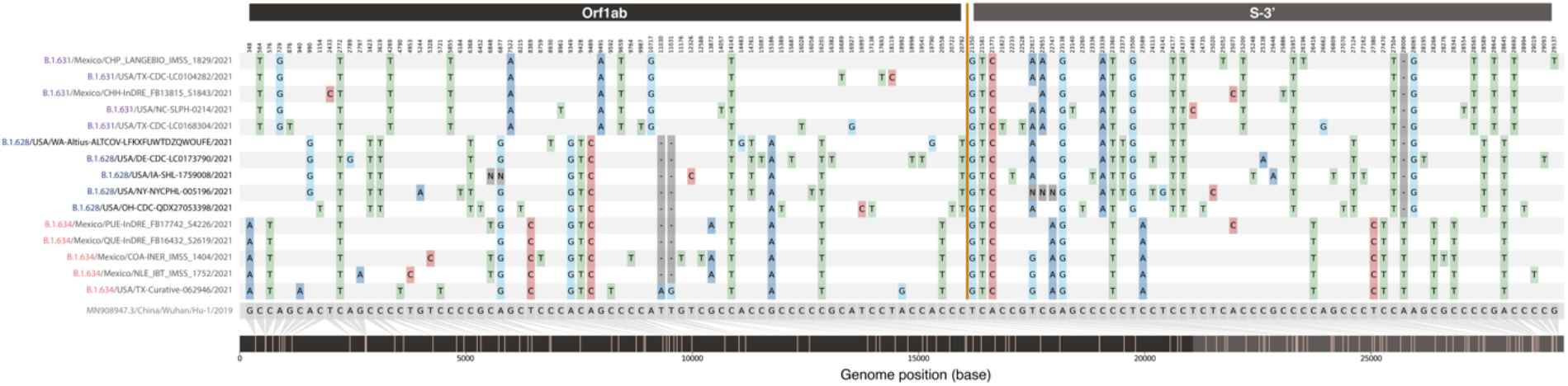
Comparison of single nucleotide polymorphisms (SNPs) between the B.1.631, B.1.628 and B.1.634 lineages. SNPs were identified in reference to the 2019 Wuhan Hu-1 reference genome (MN908947.3), shown in grey in the bottom line. Five sequences from the putative parental B.1.631 major lineage are shown on the top (purple), followed by five sequences from the putative recombinant B.1.628 major lineage on the middle (blue) and five sequences from the putative parental B.1.634 lineage at the bottom (red). Recombination point relative to the SNPs is marked by the yellow line and by different shading colours in the genome position bar (bottom of panel).

## Discussion

Genomic recombination has been widely described across sarbecoviruses in general (Boni et al., 2020) and has been identified as an important driver in the evolution of the lineage leading to the emergence of SARS-CoV-2 (Li et al., 2020; Lytras et al., 2021). More recently, recombination between the B.1.1.7 and B.1.177 lineages has been observed in the United Kingdom leading to a limited number of circulating genomes and to their designation as lineage XA, the first recombinant SARS-CoV-2 lineage under the Pango nomenclature (Jackson et al., 2021). However, no major circulating recombinant lineages spanning wider spatiotemporal distributions – across multiple countries – have been described to date. In the current work we investigate the evolutionary histories of four distinct but unusually similar SARS-CoV-2 Pango lineages circulating predominantly in the USA and Mexico and test the hypothesis that a recombination event led to the emergence of at least one of these lineages. A model can be proposed that resolves the phylogenetic incongruencies and deletion events among these lineages, in which B.1.628 major originated from a single recombination event (Fig 4). The early identification of this lineage in the USA and Mexico and its widespread circulation elsewhere represents a notable event in the COVID-19 pandemic, as no previously recognised recombinant lineages have been reported to spread across country borders and to increase in frequency at the rate observed here. Following our results, the Pango Network committee has decided that lineage B.1.628 *major* will be designated as lineage XB, making it the second recombinant lineage in the nomenclature system.

A necessary condition for the emergence of recombinant viruses is the co-circulation of its parental lineages, as viral recombination necessarily occurs during co-infection events of a single host (Simon-Loriere & Holmes, 2011). This condition is generally observed for the four lineages under investigation, predominantly detected in the USA and Mexico (where overlapping temporal distributions suggests co-circulation), as well as in other countries in Central America (Fig 1, Table 1). The substantial number of sequences and spatiotemporal distribution of B.1.628 major might normally be interpreted as evidence of an earlier emergence compared to B.1.627, B.1.631 and B.1.634. However, sampling intensity and the relative frequency of different lineages in the region require careful consideration (Kraemer et al., 2021), particularly given the considerable disparities in sampling intensity in the context of genomic surveillance (Brito et al., 2021).

B.1.627 and B.1.628 major were the first lineages to be detected and were particularly frequent in the USA (amongst these four lineages). Both increased in frequency in the USA between January and May 2021; whilst detection of B.1.627 declined to low levels by May, B.1.628 exhibited a second peak in detection in the USA in July 2021 (Fig 1; Fig S6). Mexico reported sequences, predominantly of B.1.628 major and B.1.634, from May to July (Fig 1; Fig S7), coinciding with the start of a systematic, nationwide genomic surveillance programme under the CoVi-Gen Mex Consortium on May 11 2021 (CONACYT, 2021). Given the differences in genome sampling and sequencing intensity between the two countries during the months preceding the detection of these lineages (6.3% of confirmed cases were sequenced in the USA during the last week of March 2021, compared to 1.6% for Mexico; Brito et al., 2021), it is likely that early cases of the B.1.628 major were not detected by genomic surveillance. However, the regional distribution of the lineages does suggest the recombinant lineage emerged in North America between late 2020 and early 2021.

Both minor clusters identified for B.1.628 and B.1.631 fail to display the monophyly condition which defines the Pango nomenclature (Rambaut et al., 2020). The phylogenetically distinct B.1.631 minor cluster was exclusively sampled in Turkey between late June and early August. Its spatiotemporal features and phylogenetic placement (Fig 3) and its distinct genome-wide divergence to the reference genome (Fig S2) warrants further investigation. However, its relevance in the evolution of the remaining lineages appears inconsequential.

The inferred recombination breakpoint is located near the start of the S protein reading frame, generally coinciding with previously described recombination hotspots for other coronaviruses (Sabir et al., 2016; Yang et al., 2021) and for SARS-CoV-2 (Boni et al., 2020; Jackson et al., 2021; Li et al., 2020; Lytras et al., 2021). However, the exact location of the breakpoint differs depending on the exact data set and method used. The two inferred breakpoints from our data sets (i.e., leading to lineage XB) are biologically relevant, located on the S protein signal peptide or on the RBD. A breakpoint on the S protein signal peptide would produce a viral genome in which each reading frame is inherited from one of the two parental lines in its entirety, while a breakpoint on the RBD would result in a chimeric S protein. This latter possibility remains plausible given that the breakpoint would not disrupt major functional features of the protein (such as the trimeric ACE2-binding interface) and that sequence divergence remains low between these closely related lineages. Another interesting observation is that the canonical sarbecovirus transcription regulatory sequence (TRS), a 7-nucleotide sequence that regulates the viral protein expression during cell infection (Yang et al., 2021), is located 18 nucleotides upstream of the GARD breakpoint obtained with the full data set. The TRS is also associated with viral genome replication, providing a possible mechanism driving recombination at this particular breakpoint (Yang et al., 2021). From an inferential standpoint, given the uncertainty regarding the precise location of the recombination breakpoint, exploring the individual phylogenetic tree of Orf1ab independently from the phylogeny for the remainder of the genome should adequately explain the complete evolutionary history of the lineages under investigation.

While our model for the recombinant origin of B.1.628 major reconciles the deletion and phylogenetic patterns observed in the genomic data, it still doesn’t resolve all differences between the tree topologies for the Orf1ab and S-3’ regions; for example, individual sequences with no deletions are occasionally found in clusters/lineage that are characterised by those deletions. If these sequences are correct, then that would imply repeated reversion of the deletion; this is thought to be highly unlikely as there are no known mechanisms by which a specific combination of nucleotides (i.e., the ancestral sequence) would be inserted into a site where a deletion previously occurred. The insertion of predictable short sequences has only been generally described for specific genetic elements (Sehn, 2015). We therefore conclude that these apparent reversions are more likely artifacts deriving from sequencing or assembly errors; this would be consistent with the presence of genome sequences in our data set where the deletion site falls among highly ambiguous positions (https://virological.org/t/issues-with-sars-cov-2-sequencing-data/473/12).

We find that B.1.628 major arose from a recombination event between the B.1.631 major clade and lineage B.1.634, prompting its designation as a recombinant lineage under the Pango nomenclature convention. We also recommend that the group of sequences identified here as B.1.628 minor should be revisited, and potentially designated as lineage B.1. At the time of writing, the dominance of lineage B.1.617.2 (VOC Delta) appears unchallenged at a global scale, yet the remarkable expansion of B.1.628 during early- and mid-2021 highlights the viability of a recombinant SARS-CoV-2 lineage and delineates yet another key function of active genomic surveillance programmes. Our findings also emphasise the importance of further investigating the recombination rate and potential of the virus, and of exploring the drivers of such evolutionary processes.

## Data Availability

Viral genome sequences are publicly available from GISAID (www.gisaid.org). Code used for generating analyses and figures in this study is available on GitHub (https://github.com/BernardoGG/XB_lineage_investigation).

https://github.com/BernardoGG/XB_lineage_investigation

## Acknowledgements

We would like to thank @babarlelephant for the insightful contributions to the discussions on the topic on Twitter and the cov-lineages/pango-designation GitHub account (issue #189: https://github.com/cov-lineages/pango-designation/issues/189). We also thank all laboratories and institutions involved in the generation of virus genome data publicly shared on GISAID.

## Supplementary Figures

**Figure S1.**
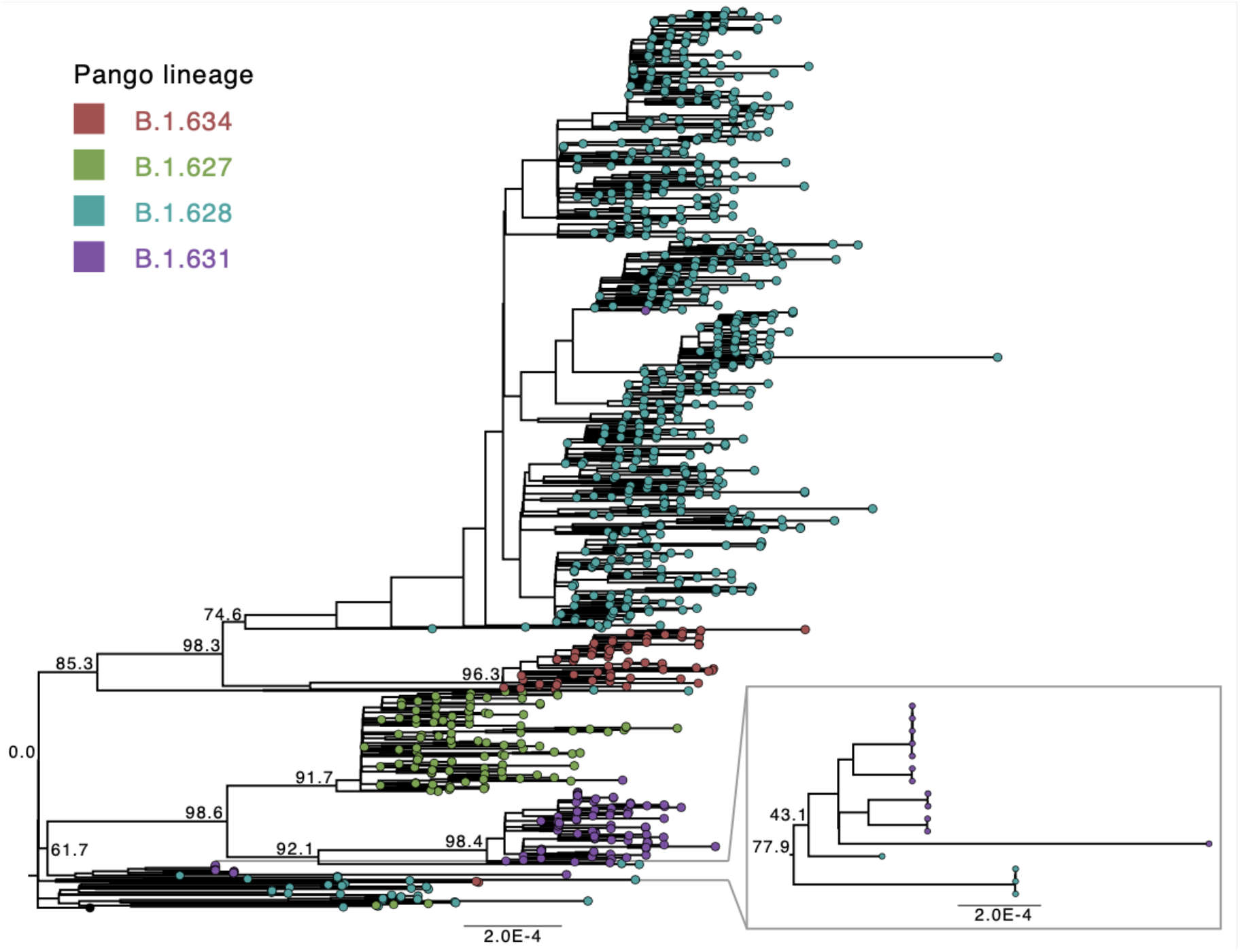
Maximum likelihood phylogenetic analyses of the complete SARS-CoV-2 genome for the four lineages. The individually designated PANGO lineage for each sequence is highlighted (the predominant lineage for sections of the tree shown), and SH-aLRT node support is shown for key lineage defining nodes on both phylogenies.

**Figure S2.**
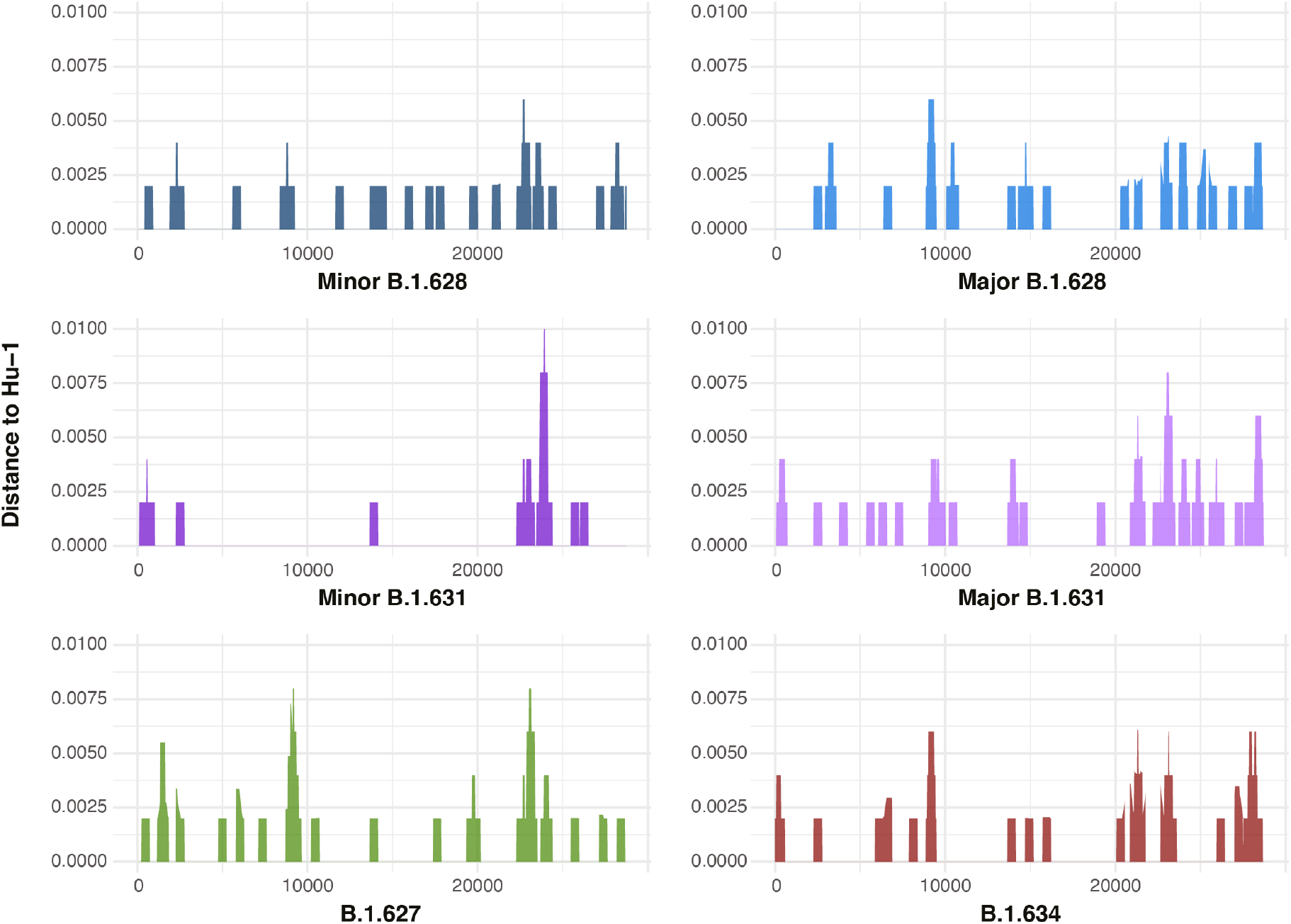
Genetic distance plots across the SARS-CoV-2 genome between the main SARS-CoV-2 phylogenetic clusters amongst the four lineages under investigation and the reference Wuhan Hu-1. Raw genetic distances between basal sequences for each of the monophyletic groups identified in the phylogenetic analyses and the 2019 Wuhan Hu-1 reference genome (MN908947.3). Distances were estimated from genomic segments of 500 nucleotides in length, overlapping over 20-nucleotide intervals.

**Figure S3.**
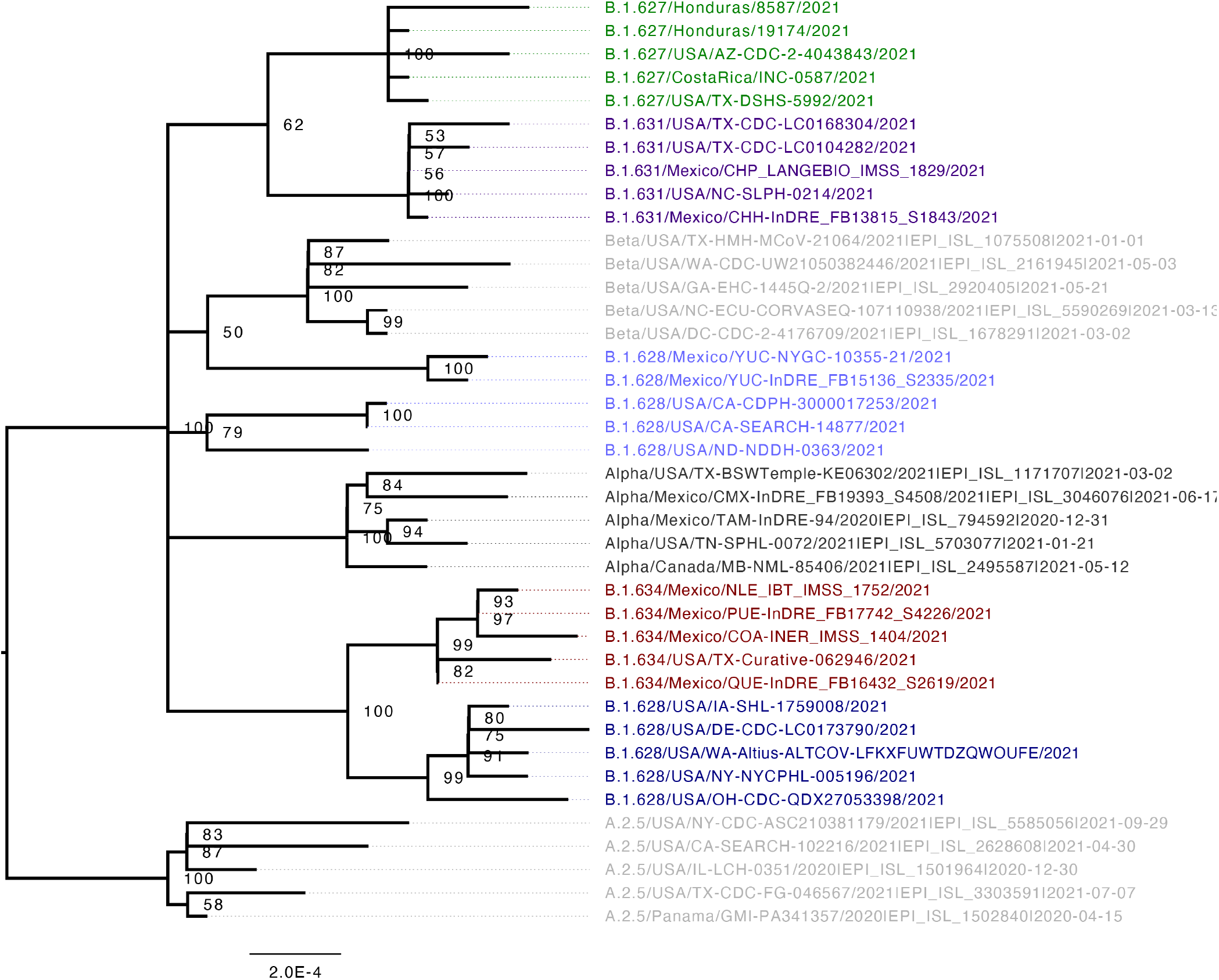
Maximum likelihood phylogenetic tree for the Orf1ab genome segment of a selection of sequences of lineages under investigation B.1.627 (green), B.1.628 minor (light blue), B.1.628 major (dark blue), B.1.631 (purple) and B.1.634 (red) in relation to B.1.1.7 (VOC Alpha, dark grey) and B.1.351 (VOC Beta, fark grey). Node support is shown from 1000 bootstrap replicates, nodes with support <50% are collapsed into polytomies. The tree is rooted in reference to lineage A.2.5.

**Figure S4.**
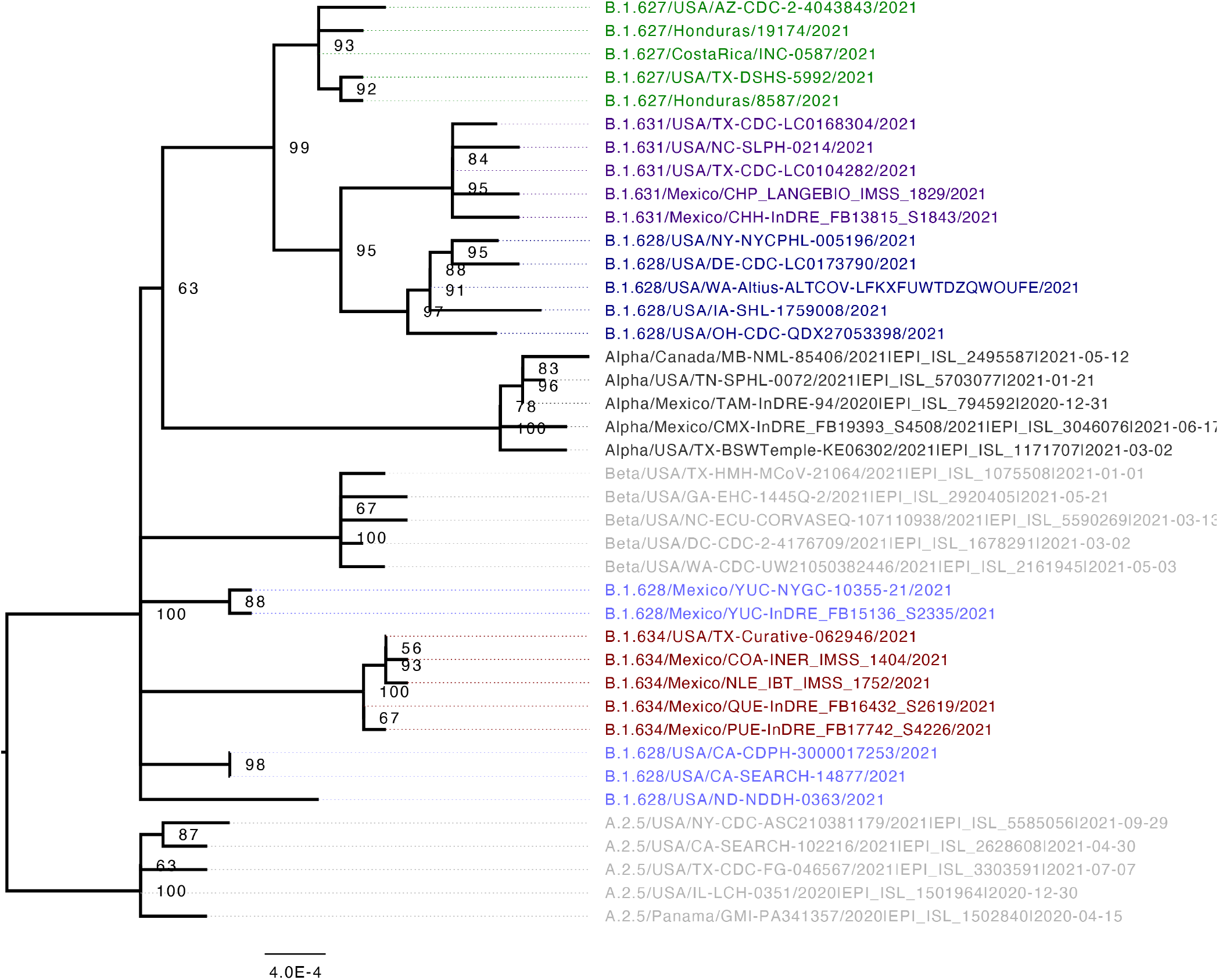
Maximum likelihood phylogenetic tree for the S-3’ genome segment of a selection of sequences of lineages under investigation B.1.627 (green), B.1.628 minor (light blue), B.1.628 major (dark blue), B.1.631 (purple) and B.1.634 (red) in relation to B.1.1.7 (VOC Alpha, dark grey) and B.1.351 (VOC Beta, fark grey). Node support is shown from 1000 bootstrap replicates, nodes with support <50% are collapsed into polytomies. The tree is rooted in reference to lineage A.2.5.

**Figure S5.**
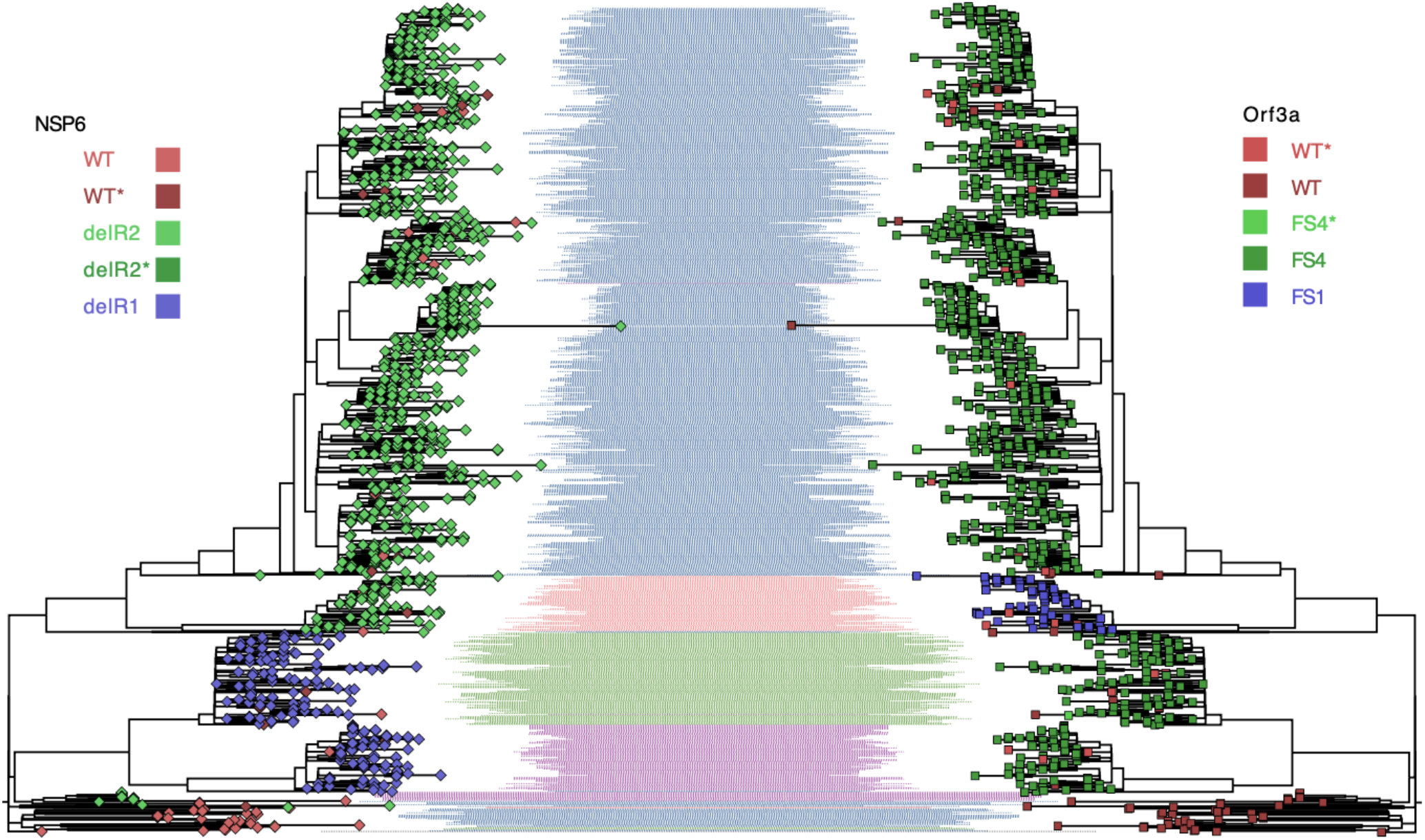
Deletions on the Orf1ab (NSP6; diamonds) and Orf3a (rectangles) loci mapped to the maximum likelihood phylogeny of the complete genome for the four lineages under investigation. Major lineage designations are shown in coloured shading: B.1.627 (green), B.1.628 (blue), B.1.631 (purple) and B.1.634 (red). Darker colours assigned to tips correspond to sequences where the marked deletion occurs amongst ambiguous sites (Ns).

**Figure S6.**
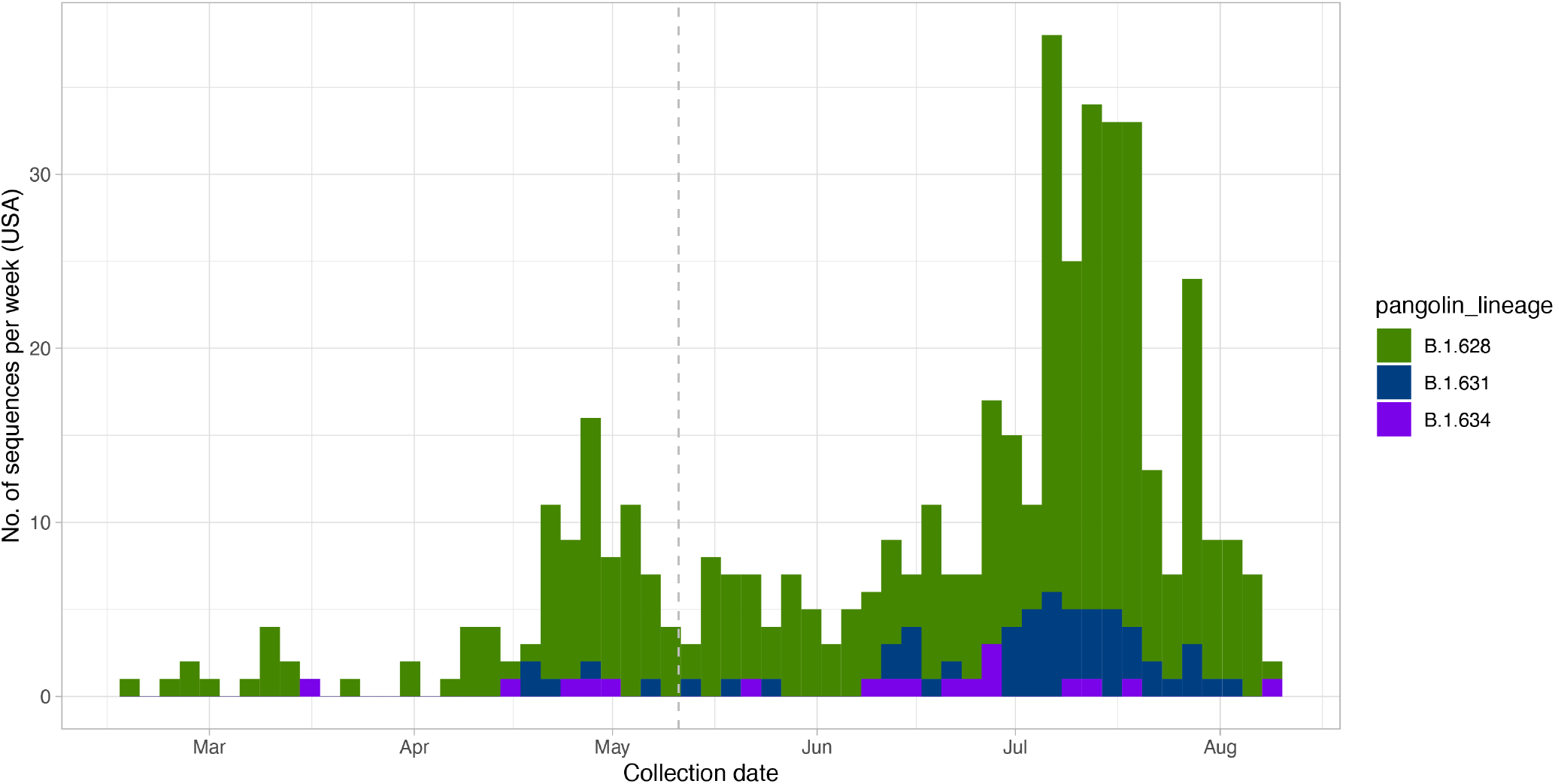
Sequence sampling from the B.1.628 major, B.1.631 and B.1.634 lineages in the USA. Sequences shown here correspond only to the sequences included in the phylogenetic analyses (i.e. <10% ambiguities in the genome sequence, >90% genome coverage). Dotted line shows the starting date for the systematic genomic surveillance work performed in Mexico by the CoVi-Gen Mex Consortium.

**Figure S7.**
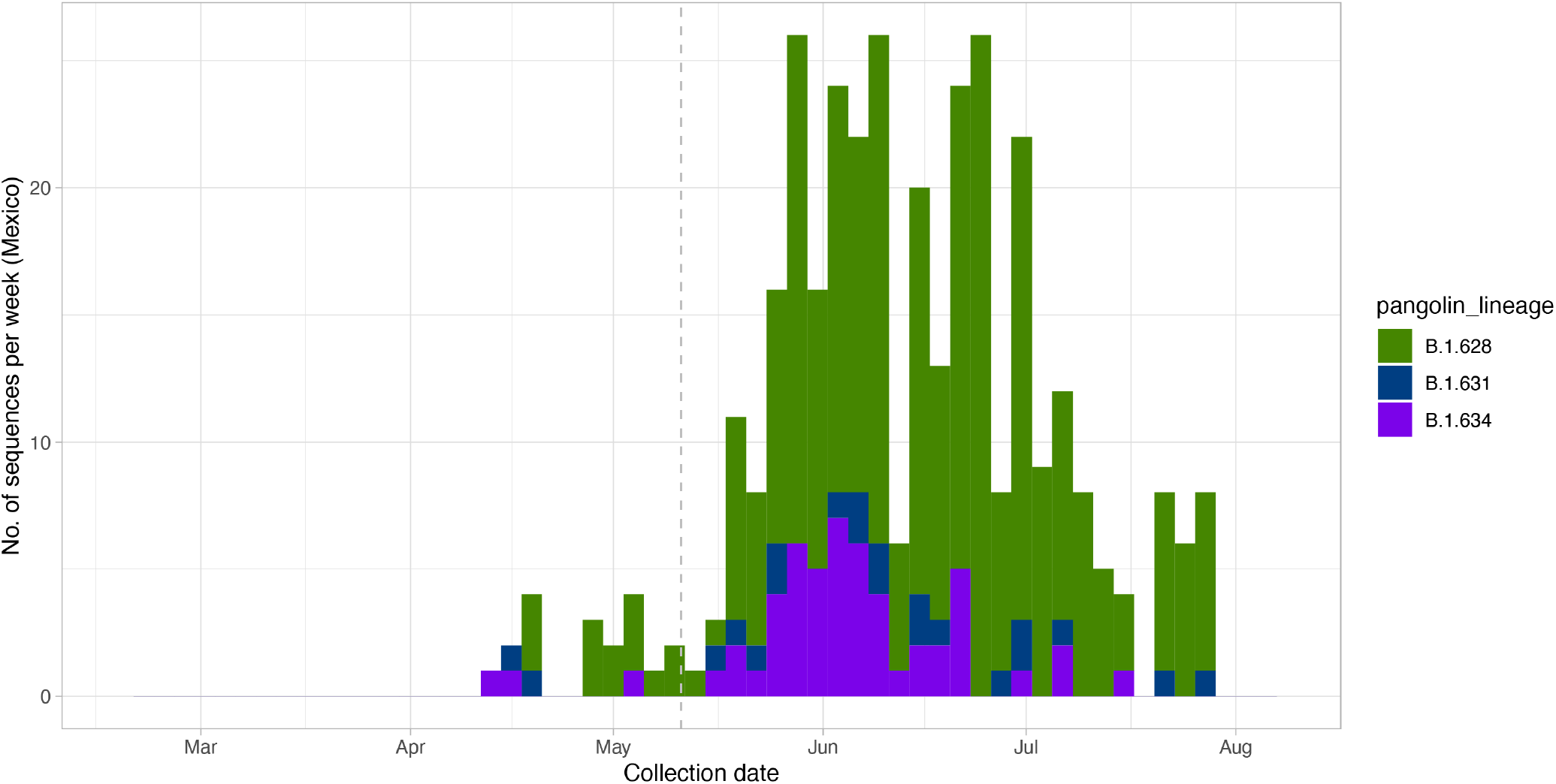
Sequence sampling from the B.1.628 major, B.1.631 and B.1.634 lineages in Mexico. Sequences shown here correspond only to the sequences included in the phylogenetic analyses (i.e. <10% ambiguities in the genome sequence, >90% genome coverage). Dotted line shows the starting date for the systematic genomic surveillance work performed in Mexico by the CoVi-Gen Mex Consortium.

